# Probability of aerosol transmission of SARS-CoV-2

**DOI:** 10.1101/2020.07.16.20155572

**Authors:** Scott H. Smith, G. Aernout Somsen, Cees van Rijn, Stefan Kooij, Lia van der Hoek, Reinout A. Bem, Daniel Bonn

**Affiliations:** Van der Waals-Zeeman Institute, Institute of Physics, University of Amsterdam, Amsterdam, The Netherlands; Cardiology Centers of the Netherlands, Amsterdam, The Netherlands; Laboratory of Experimental Virology, Department of Medical Microbiology, Amsterdam UMC, Location AMC, University of Amsterdam, Amsterdam, the Netherlands; Department of Pediatric Intensive Care, Emma Children’s Hospital, Amsterdam University Medical Centers, location AMC, Amsterdam, The Netherlands

**Author notes:** <u>Correspondence:</u> Prof. Dr. Daniel Bonn, *Institute of Physics, University of Amsterdam, Science Park 904, 1098XH Amsterdam, The Netherlands,.

**Keywords:** SARS-CoV-2, COVID-19, aerosol, viral transmission, infection, respiratory droplets

## Abstract

Transmission of SARS-CoV-2 leading to COVID-19 occurs through exhaled respiratory droplets from infected humans. Currently, however, there is much controversy over whether respiratory aerosol microdroplets play an important role as a route of transmission. By measuring and modeling the dynamics of exhaled respiratory droplets we can assess the relative contribution of aerosols in the spreading of SARS-CoV-2. We measure size distribution, total numbers and volumes of respiratory droplets, including aerosols, by speaking and coughing from healthy subjects. Dynamic modelling of exhaled respiratory droplets allows to account for aerosol persistence times in confined public spaces. The probability of infection by inhalation of aerosols when breathing in the same space can then be estimated using current estimates of viral load and infectivity of SARS-CoV-2. In line with the current known reproduction numbers, our study of transmission of SARS-CoV-2 suggests that aerosol transmission is an inefficient route, in particular from non or mildly symptomatic individuals.

## Introduction

Respiratory droplets form the most important carrier of SARS-CoV-2 virions, and may infect humans by direct inhalation or indirectly through hand or object contact. During the current COVID-19 pandemic, numerous explosive local outbreaks, so called super-spreading events, in public spaces or health care settings have raised the concern of aerosol transmission of SARS-CoV-2. Aerosols, or microdroplets, are formed and exhaled during loud speaking, singing, sneezing and coughing. As infected persons (initially) may have none or mild symptoms, an aerosol transmission route of SARS-CoV-2 may have tremendous impact on health care strategies to prevent the spreading of COVID-19 in public spaces. Importantly, SARS-CoV-2 viral particles have been detected in microdroplets which may spread in exhaled air during breathing, talking, singing, sneezing, or coughing by an infected individual.^1-3^ Microdroplets form aerosol clouds, which have a relatively long airborne time,^4^ and may thus pose an important threat to community spread of COVID-19. However, to what extent microdroplets in practice result in infections with the SARS-CoV-2 virus, remains a topic of intense debate.^5-9^ Next to virus and host factors, this type of viral transmission through aerosols depends strongly on droplet properties and behavior.^10,11^ In order to aid the development of effective preventive strategies for SARS-CoV-2 transmission, in this study we measure and model respiratory droplet physics to predict the importance of community SARS-CoV-2 transmission by the aerosol route.

## Results and discussion

We measure size distributions of droplets in aerosols released when speaking or coughing using laser diffraction (Malvern Spraytech®), and consistently find a double-peaked drop size distribution for coughing, and a single-peak drop size distribution for speech, which can be described by a distribution corresponding to a normal liquid spraying process,^11^ as shown in Figure 1. A previous study,^2^ showed that age, sex, weight, height, or corporal mass have no statistically significant effect on the aerosol composition in terms of size and number of droplets. We tested 7 healthy volunteers (5 male, 2 female), and found that the variability in drop production by coughing between the different emitters was relatively small, except for one person, who produced 17 times more liquid volume than the others. It has been suggested that if such a person would be infected with SARS-CoV-2, he or she could become a so-called “super-spreader” due to the high number of droplets emitted,^2,3^ although droplet dynamics in symptomatic SARS-CoV-2 infected humans are unclear.

**Figure 1.**
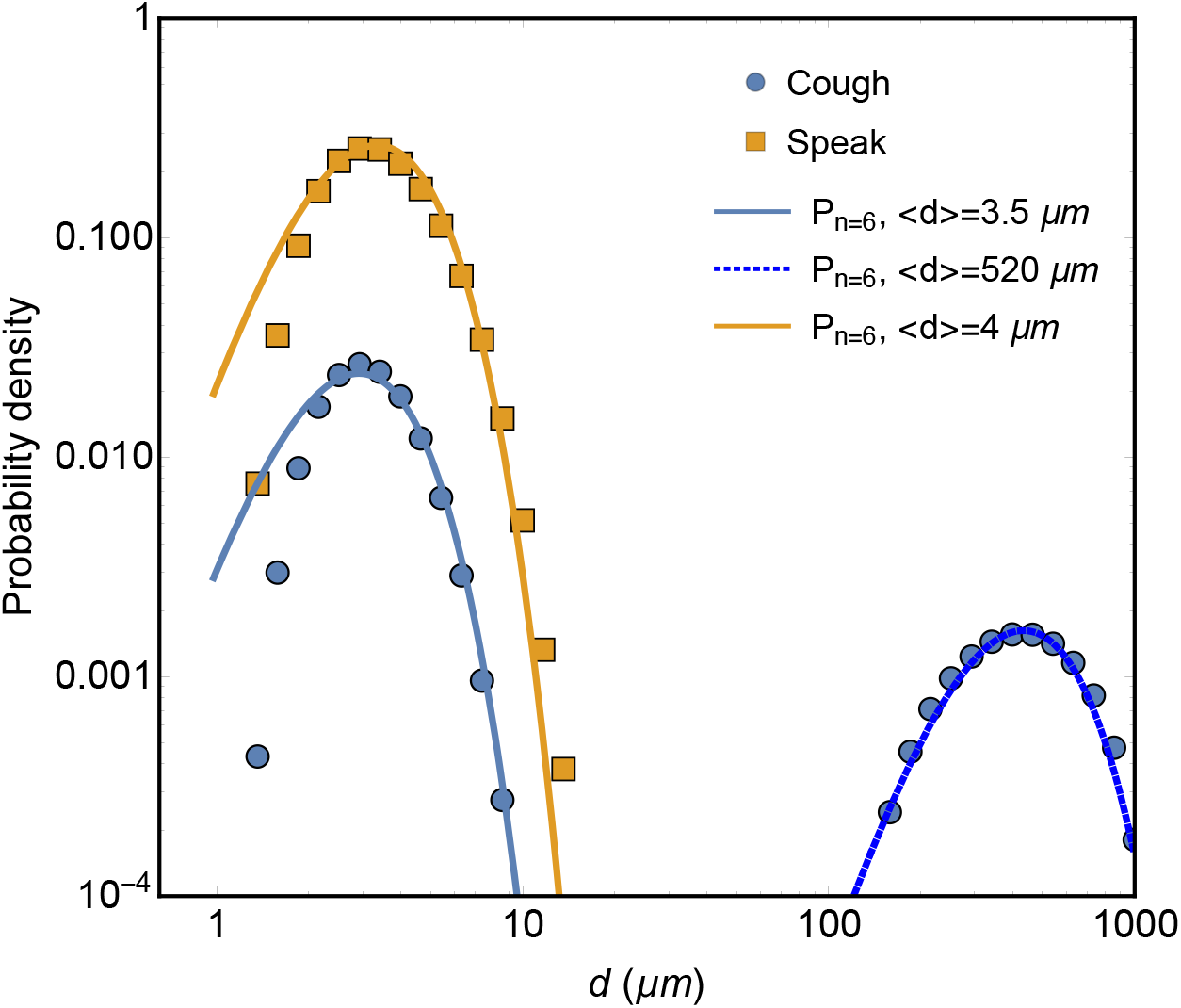
Measured drop size distributions of droplets produced when coughing (circles) and speaking (squares). Solid lines are fits with Gamma distributions.^11^

Using a precision balance, the volumes of saliva/mucus produced by the high emitter when coughing or speaking into a small plastic bag were measured by weighing before and after a single cough or saying “Stay Healthy” for 10 times.^12^ Averaging over 20 experiments, we find that a single cough yields a liquid weight of 0.07 ± 0.05 gram, whereas speaking ten times produces a weight of 0.003 ± 0.001 gram.

For coughing, the volumetric distribution measured using laser diffraction shows that on average 98 ± 1% of the volume of the spray is contained in the large drops (100-1000 µm). For the small aerosol droplets, this amounts to ∼20 million microdroplets produced in a single cough and ∼7 million for speech. For COVID-19, thus from symptomatic patients, viral RNA load in undiluted oral fluid or sputum has been found to be 10^4^ - 10^6^ copies per milliliter.^13-16^ Although temporal changes in viral load may be correlated to COVID-19 severity, with very high viral loads up to ∼10^11^ copies per milliliter,^14^ this dynamic association so far has not been definitely established. As such, following the Ref.^13^ to avoid underestimation, we used a number of 7×10^6^copies per milliliter in respiratory samples in our primary analysis. The total number of virus particles present in the total volume of only the microdroplets is then 10^4^, implying that only 1 in 2000 aerosol droplets contains a virus particle.

The persistence of these aerosol droplets in the air is of the greatest concern regarding community transmission of SARS-CoV-2 in public spaces. This airborne time is governed by evaporation and gravity-driven sedimentation towards the floor. The latter can be explained by balancing the forces of gravity (*F=mg*) and air drag (*F=6πηRU*, with *η* the air viscosity, *R* the droplet radius, and *U* the falling velocity), from which it follows that a typical small droplet will take nine minutes to reach the ground. This time will even increase by the evaporation of the liquid phase of the droplet. Sputum droplets are known to consist for 1-10% of their volume of solid solutes.^17^ Consequently, they will not evaporate completely but leave a ‘solid’ core residue. For microdroplets smaller than 10 µm, the contraction to the solid core having half of the original droplet size (i.e., ∼10% of the initial volume) happens within a second in quiescent air with a relative humidity of 50%,^17^ and a droplet half the size stays airborne four times longer.

A laser light sheet was used to track microdroplets similar to those produced by coughing and speaking. To mimic small respiratory droplets, we used a mixture of 1% glycerol and 99% ethanol; within a second, the ethanol evaporates, yielding small non-evaporating droplets of glycerol with a median mass aerodynamic diameter (MMAD) of 5 ± 3 µm, similar to the microdroplets produced by for coughing or speaking. The number of drops passing through the laser sheet suspended in the center of our 2×2×2 m^3^ experimental chamber was analyzed using an algorithm that detects the illuminations caused by the droplets. Typical results are shown in Figure 2 and capture the reduction in number of droplets over time due to coupled effects of sedimentation, horizontal displacement, and evaporation.

**Figure 2.**
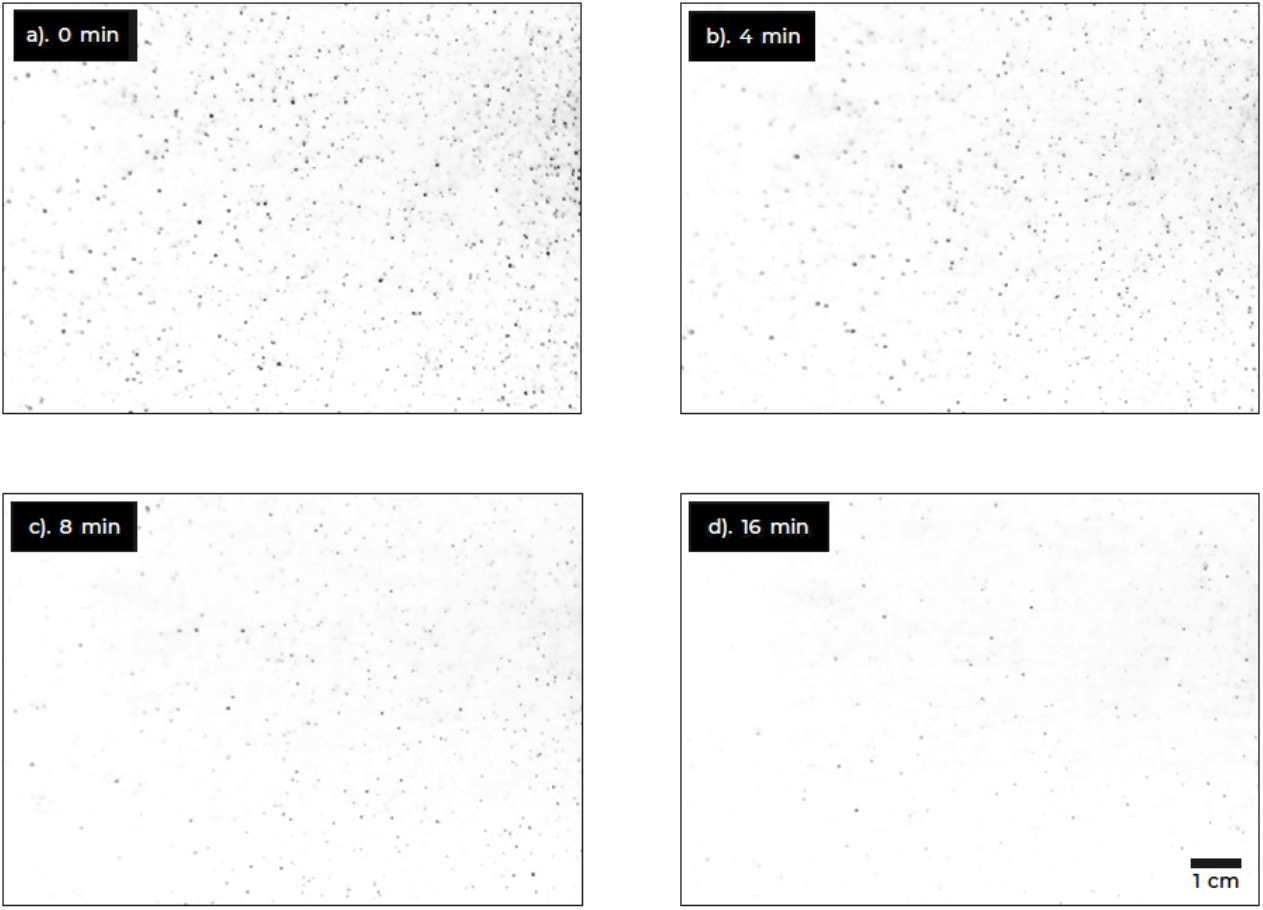
Laser-illuminated aerosol droplets at different times after initial spraying. Initially (**a**), droplets have a maximum sedimentation velocity of about 2 cm/sec, corresponding to droplets of about 25 µm in diameter. In the 16 min frame (**d**), the fastest moving droplet has a sedimentation velocity of at most 1 mm/sec, corresponding to a droplet of about 4-5 µm in diameter.

If these aerosol droplets are a vector of transmission for the SARS-CoV-2 virus, how the number of droplets decreases as a function of time will have a significant influence on the potential airborne transmission of SARS-CoV-2. To predict the evolution in number of microdroplets, the evaporation and sedimentation can be accounted for to calculate the number of airborne aerosol particles (see the Supplementary Materials for details of the calculation). Figure 3 compares our predictions for droplet persistence results with our own results and those reported by others.^3^ It shows that the model accurately captures the exponential decline in the number of droplets over time for both experiments and suggests the decline is, to a small extent, influenced by the evaporation of the droplets (i.e., the relative humidity of the environment) but dominated by the sedimentation. Additionally, from Figure 3, it can be concluded that the time to half the original number of droplets in the system (i.e., the half-life) is between 5.5 and 7 min.

**Figure 3.**
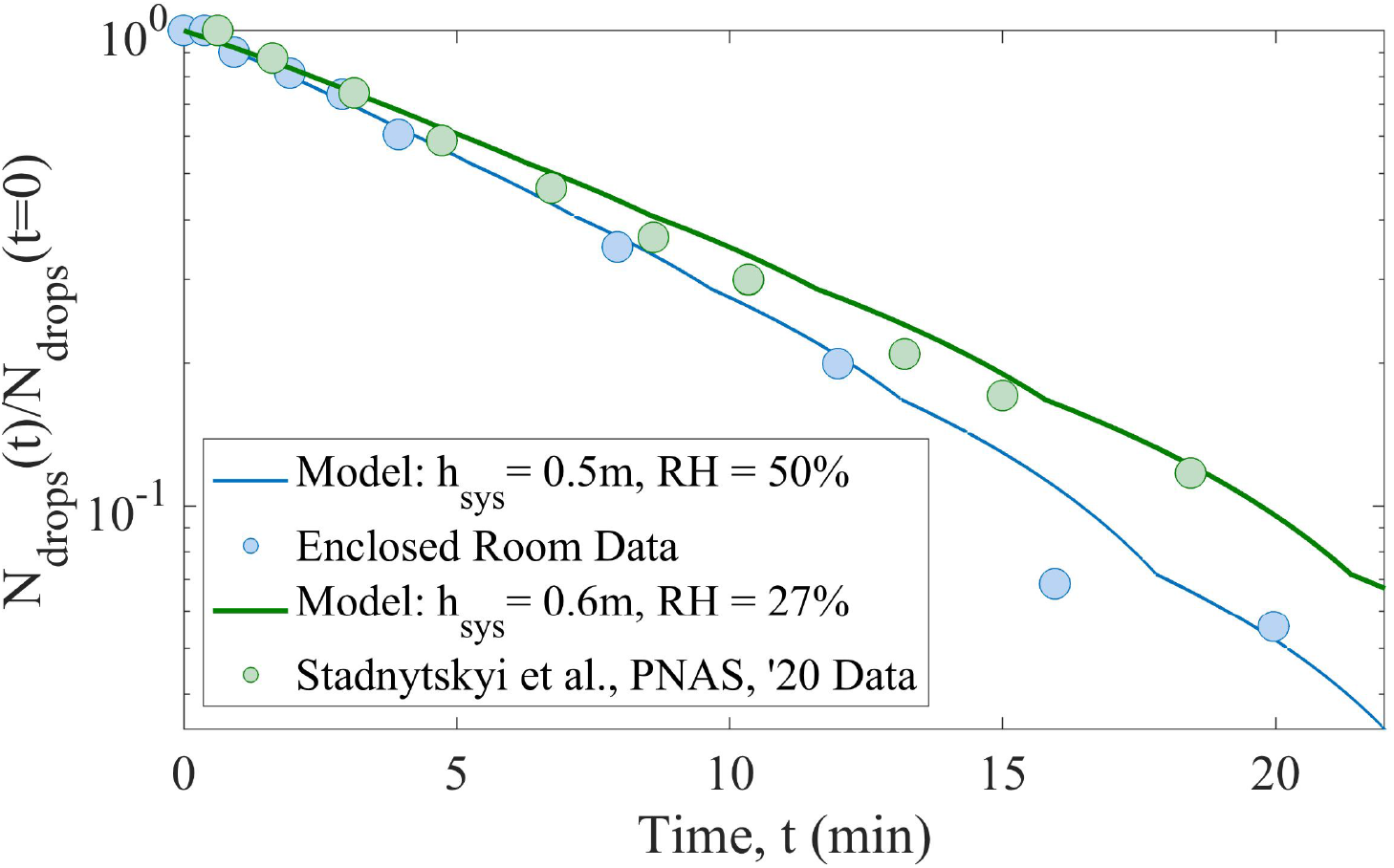
Normalized number of droplets as a function of time as determined experimentally (blue circles) compared to the data of Ref.^3^ (green circles). Solid lines are model fits for both sets of data, with input parameters relative humidity (RH) and sedimentation height (*h*_*sys*_).

This then allows us to estimate how many virus particles one would inhale while inside a room where an infected person coughed a single time. The highest probability of infection occurs when a person enters a poorly ventilated and small space where a high emitter has just coughed, and inhales virus-carrying droplets. We model coughing in our 2×2×2 m^3^ unventilated space that .could represent e.g. a restroom. The drop production by coughing was found to be very similar for 6 out of the 7 emitters. We find peak values of 1.18 ± 0.09 × 10^3^ pixels that light up in the field of view of our laser sheet (21×31 cm^2^). This directly corresponds to the volume of emitted droplets^3^; the high emitter produced 1.68 ± 0.20 × 10^4^ lit up pixels, more than an order of magnitude larger. Based on these numbers and the earlier measured volume and drop size, we can calculate the amount of virus inhaled by a person entering and staying in the same room where an infected person produced the droplets, as a function of entrance delay and residence time. As detailed above, the calculation assumes a viral load of 7 × 10^6^ copies per milliliter of saliva.^13^ We also assume a single inhalation volume of 0.0005 m^3^ (tidal volume 6 ml per kg body weight for an adult man) and a normal respiratory rate of ∼16 inhalations/min.^18^ In Figure 4, we compare the results for the high emitter with those for a regular (low) emitter on the basis of the amount of light scattered from droplets produced by a single cough.

**Figure 4.**
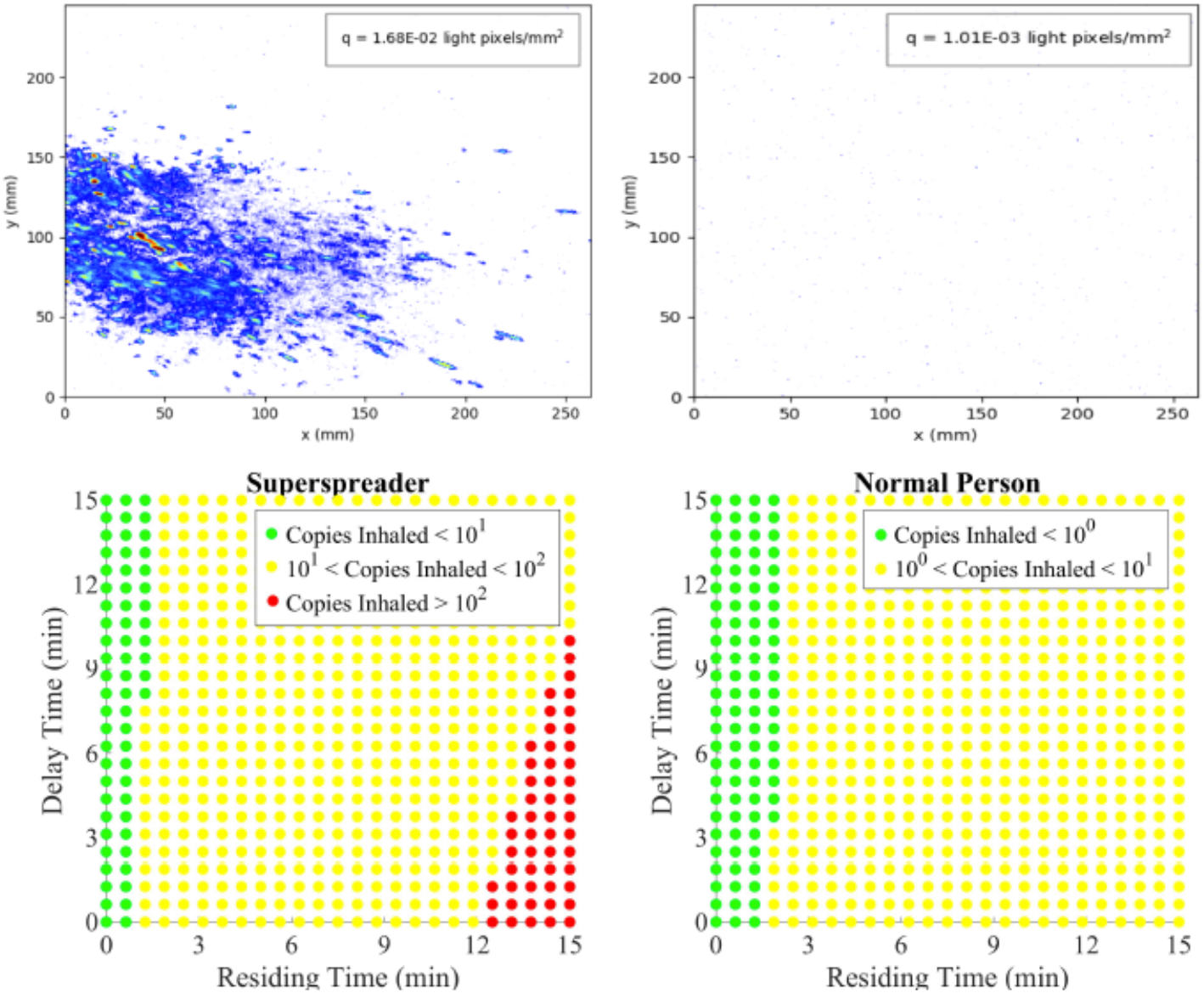
Instantaneous pictures of the droplets produced by coughs of a high (**a**) and a normal emitter (**b**) as detected with laser sheet imaging. The cough volumes allow us to estimate the number of inhaled virus particles as a function of (i) the delay between the cough and a healthy person entering the room, and (ii) the time the healthy person spends in the room (**c**,**d**).

The number of virus particles needed to infect a single individual, *N*_*inf*_, needs to be taken into account to translate these findings into risk of infection. This obviously also depends on factors such as the vulnerability / susceptibility of the host, yet as detailed in Ref.^19^, the respiratory infectivity for SARS-CoV-2 is not yet well known. In the absence of data on SARS-CoV-2, the most reasonable assumption is that the critical number of virus particles to cause infection is comparable to that for other Coronaviruses, including SARS-CoV-1, and influenza virus. In that case, *N*_*inf*_ ∼100-1000, which corresponds to ∼10-100 PFU.^19-21^. If we adopt a conservative approach and assume the upper limit of this range (*N*_*inf*_ ∼100), we find that our unventilated 2×2×2 m^3^ space contaminated by a single cough is safe for residing times of less than 12 minutes due to the low virus content of the aerosol particles. Additionally, the maximal number of inhaled viral copies by a person entering the room after the high emitter has coughed is ∼120 ± 60, where the error margin comes from variation in relative volume of small and large drops produced by a cough. If the infected person is a regular emitter, the probability of infecting the next visitor of the confined space by means of a single cough for any delay or residence time is therefore very low. For speech, due to the low volumes emitted, this probability is even smaller. Our small non-ventilated room is also a ‘worst-case’: in better ventilated, large rooms, the aerosols become diluted very rapidly.^4^

## Conclusion

Our dynamic modelling of transmission of SARS-CoV-2 in confined spaces shows that aerosol transmission seems an inefficient route, in particular from non or mildly symptomatic individuals. The large droplets that are believed to be responsible for direct and nosocomial infections may contain about 500 virus particles per droplet and are thus likely the most important route in a mixed transmission model.

A limitation to our study is that we cannot easily take changes in virus viability inside microdroplets into account, which depend on the local microenvironment of the aerosol gas clouds as produced under different circumstances.^22^ However, viable SARS-CoV-2 in aerosols can be found after several hours,^23^ and as such this limitation will not likely affect our main conclusion. Importantly, our results do not completely rule out aerosol transmission. It is likely that large numbers of aerosol drops, produced by continuous coughing, speaking, singing, or by certain types of aerosol-generating medical interventions, can still result in transmission, in particular in spaces with poor ventilation.^4^ Our model explains the rather low reproduction number of SARS-CoV-2 in environments where social distancing is practiced compared to the reproduction numbers of other “true” airborne pathogens.^10, 24, 25^

## Materials and methods

Size distributions of droplets from aerosols released when speaking or coughing were measured using laser diffraction employing a Malvern Spraytech® with a 300mm lens. In this configuration drop sizes between 0.2 μm and 2mm can be measured. Speaking and coughing is done directly into the laser beam and data acquisition is done in the ‘fast acquisition’ mode so that there is no dead time and the drop size distribution is measured before evaporation.

## Data Availability

Not applicable.

## Acknowledgements

No medical writer or editor was involved.

## Author contributions

SHS contributed to the design, analysis and revision of the manuscript, GAS contributed to the inception, design, interpretation and revision of the manuscript, CR contributed to the inception, design, and analysis, SK contributed to the analysis, LH contributed to the interpretation and revision of the manuscript, RAB contributed to the inception, interpretation and revision of the manuscript, DB contributed to the inception, design, analysis and prepared the first draft of the manuscript.

## Test Subjects

Part of this study included measurements from subjects, which was approved by a local ethical committee (AMC 2020_098/NL73585.018.20).

